# The drop in reported invasive pneumococcal disease among adults during the first COVID-19 wave in the Netherlands explained

**DOI:** 10.1101/2021.06.29.21259494

**Authors:** Kirsten Dirkx, Bert Mulder, Annelies Post, Martijn Rutten, Caroline Swanink, Heiman Wertheim, Amelieke Cremers

## Abstract

*Streptococcus pneumoniae* is the main bacterial pathogen causing respiratory infections. Since the COVID-19 pandemic emerged, less pneumococcal disease was identified by surveillance systems around the world. Measures to prevent transmission of SARS-CoV-2 also reduce transmission of pneumococci, but this would gradually lead to lower disease rates. Here, we explore additional factors that have contributed to the instant drop in pneumococcal disease cases captured in surveillance. Our observations on referral practices and other impediments to diagnostic testing indicate that residual IPD has likely occurred but remained undetected by conventional hospital-based surveillance. Depending on setting, we discuss alternative monitoring strategies that could improve sight on pneumococcal disease dynamics.

## Background

The bacterium *Streptococcus pneumoniae* (the pneumococcus) is the leading cause of community acquired pneumonia and meningitis worldwide [1, 2]. Children under five and adults above the age of 65 are most at risk for pneumococcal infections [3]. Mortality among hospitalized patients is 10-15%. Lower respiratory tract infections are the major cause of sepsis [4-6] and account for the largest proportion of hospitalizations due to infectious diseases [7], already before the COVID-19 pandemic. In less than 10% of all pneumococcal infections in the Netherlands, the pathogen is identified by culture of a normally sterile site like blood or cerebrospinal fluid, which is called invasive pneumococcal disease (IPD) [8]. This percentage is likely an underestimation of the prevalence of bacteremia among adults with a pneumococcal infection. Collection of blood cultures is often limited to patients who present at the hospital with fever and severe respiratory disease. However, of captured pneumococcal bacteremias, 29% occurred in patients with normal body temperature and 30% was classified as mild pneumonia (this study). Furthermore, pneumococci are fragile bacteria whose culture is easily disturbed, also by prior antibiotics use [9, 10].

Because of the introduction of pediatric pneumococcal vaccination programs, in many countries the incidence of IPD is under surveillance. During the first COVID-19 wave in the Netherlands, an 80% drop in IPD was reported by the Netherlands Reference Laboratory of Bacterial Meningitis (NRLBM) [11]. In three hospitals in the East of the Netherlands we recognized this trend and additionally noted a high mortality among adult IPD patients. To investigate these observations, we compared the IPD cases during the first COVID-19 wave to the corresponding months in the 5 years preceding. We considered changes in pneumococcal exposition, delayed or waived referral to the hospital, hampered capacity to diagnose IPD (including prior antibiotics use), and potential adjustments in standards of hospital care as possible explanatory factors. Understanding what sample of adult pneumococcal infections has been captured in the IPD registration, is of importance for surveillance purposes as well as for individual case management in subsequent COVID-19 waves.

### Hospital based survey

During the first COVID-19 wave in March, April, and May 2020 a total of 13 adults with pneumococcal bacteremia were hospitalized in the 3 participating hospitals, compared to 32 ± 6 (mean ± SD) cases during the corresponding months in the five years preceding (Figure 1A). Among these 13 cases 30-day mortality was 30.8% compared to 9.9% (16 out of 161) in earlier years (p = 0.046) (Figure 1B).

**Figure 1:**
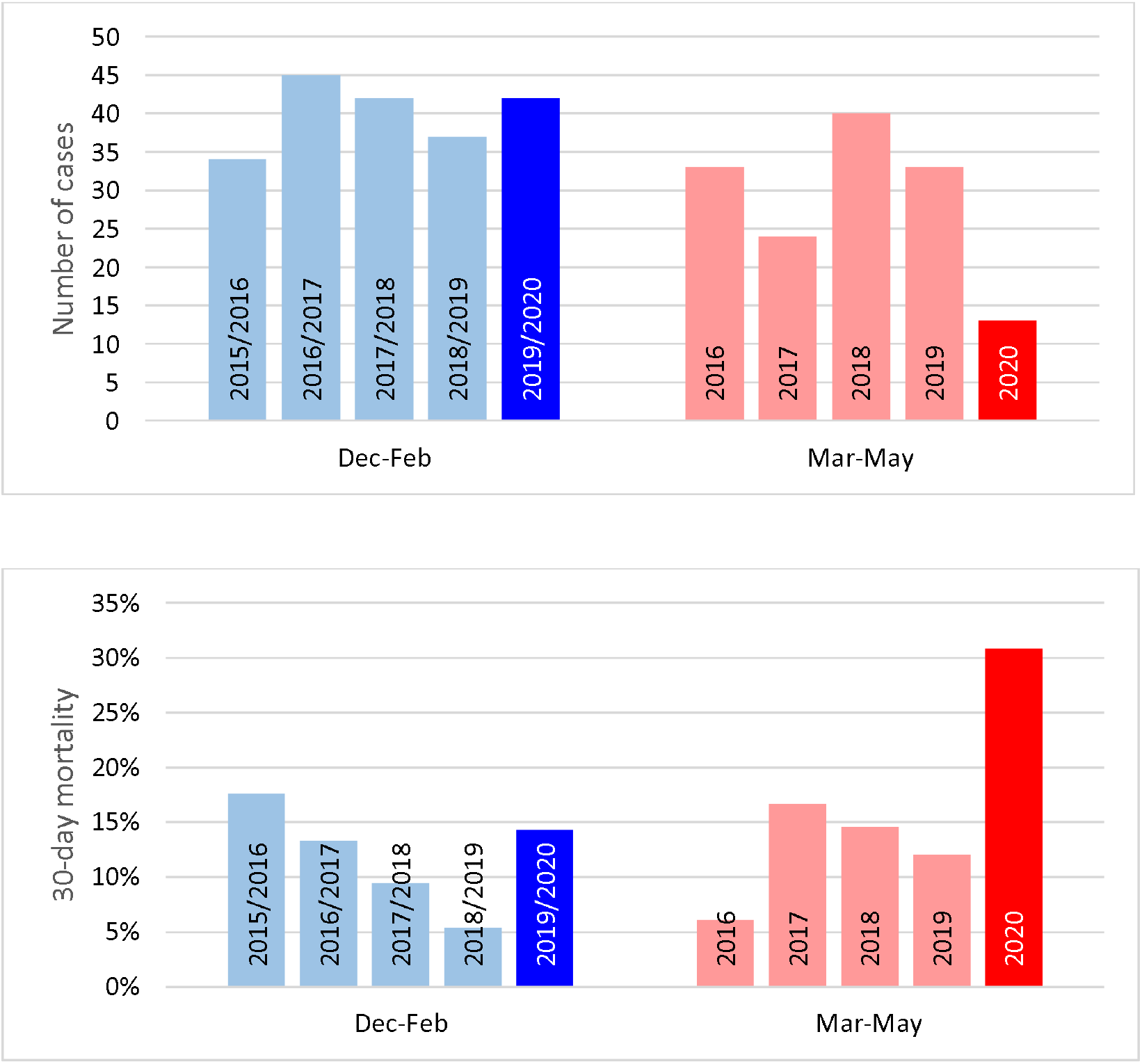
Annual bacteremic adult IPD cases hospitalized at the 3 study centers together. Cases are grouped by their identification date in 3-month time periods: December to February or March to May. Displayed are number of cases (panel A) and 30-day mortality (panel B).

### Exposition and ongoing disease burden

Nasopharyngeal pneumococcal carriage is most prevalent among children under the age of 5 who are the main source of circulating *S. pneumoniae* [12, 13]. By contrast, pneumococcal colonization is only occasionally detected among older adults [14]. The timeline in Figure 2 describes measures taken by the national government to control the COVID-19 outbreak, in relation to the number of IPD cases identified at the three participating hospitals. During the first COVID-19 wave daycare and primary schools were closed from mid-March until early May. Social distancing was advocated and included maintaining a 1.5-meter distance, restrictions on crowding, and the elderly were advised to withhold from interaction with their grandchildren. It is likely that exposure to pneumococcal carriers decreased among elderly, which can partly explain the drop in observed IPD cases.

**Figure 2:**
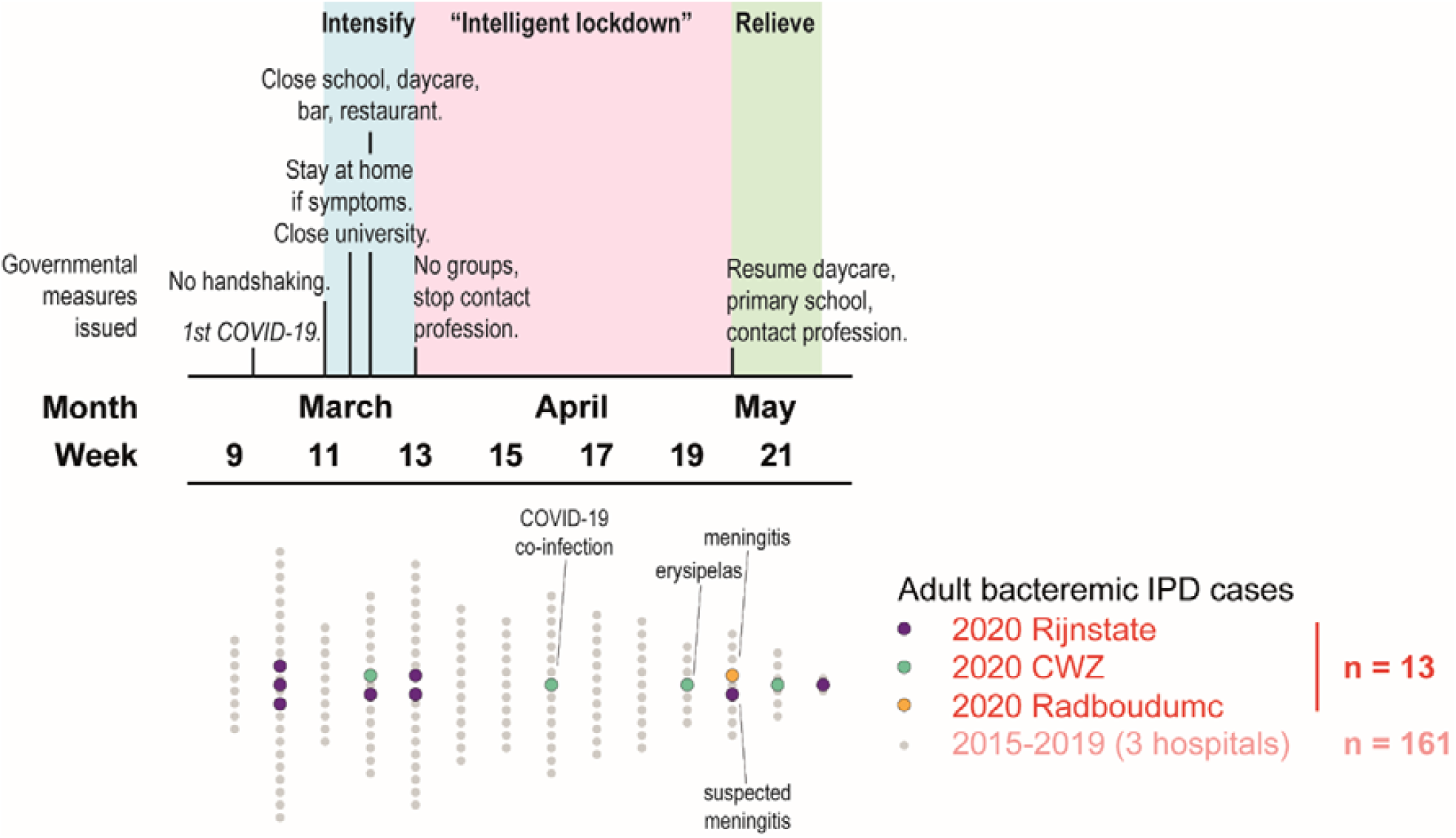
Timeline of governmental measures issued (upper graph) and bacteremic adult IPD cases hospitalized (lower graph) during the first COVID-19 wave in 2020 in the Netherlands.

However, a recent study showed that 32% of healthy volunteers of 50-84 years old were still colonized by *S. pneumoniae* at four weeks after experimental challenge [15]. This suggests that into the COVID-19 pandemic, the elderly who were already colonized by *S. pneumoniae* were still at risk for pneumococcal infection despite social distancing measures. Also, for the IPD cases in May it is not excluded that they have acquired pneumococci from external pneumococcal carriers long before developing disease. The presence of high avidity serotype-specific antibodies in elderly with IPD indicates that weeks of antibody maturation can take place prior to actual infection [16]. In addition, by molecular methods *S. pneumoniae* is still being detected in nasopharyngeal samples after cultures have become negative, and endogenous pneumococcal low-density carriage can again increase over time [17].

In the Netherlands the prescription of amoxicillin by general practitioners (GPs) is fairly always intended as the first line treatment of a bacterial pneumonia. Data from GPs’ pharmacists in the study area demonstrate that the prescription of amoxicillin during the first COVID-19 wave decreased overall by 25%, but the level of prescriptions sustained among adults 21-40 years old and those 75 and over (Figure 3). This could be due to persistent occurrence of pneumococcal respiratory infections in these age categories. At the same time, age over 75 has been an important reason not to withhold antibiotics as it is a risk factor for bacterial pneumonia, which may be hard to differentiate from COVID-19 according to the Dutch College of General Practitioners [18].

**Figure 3:**
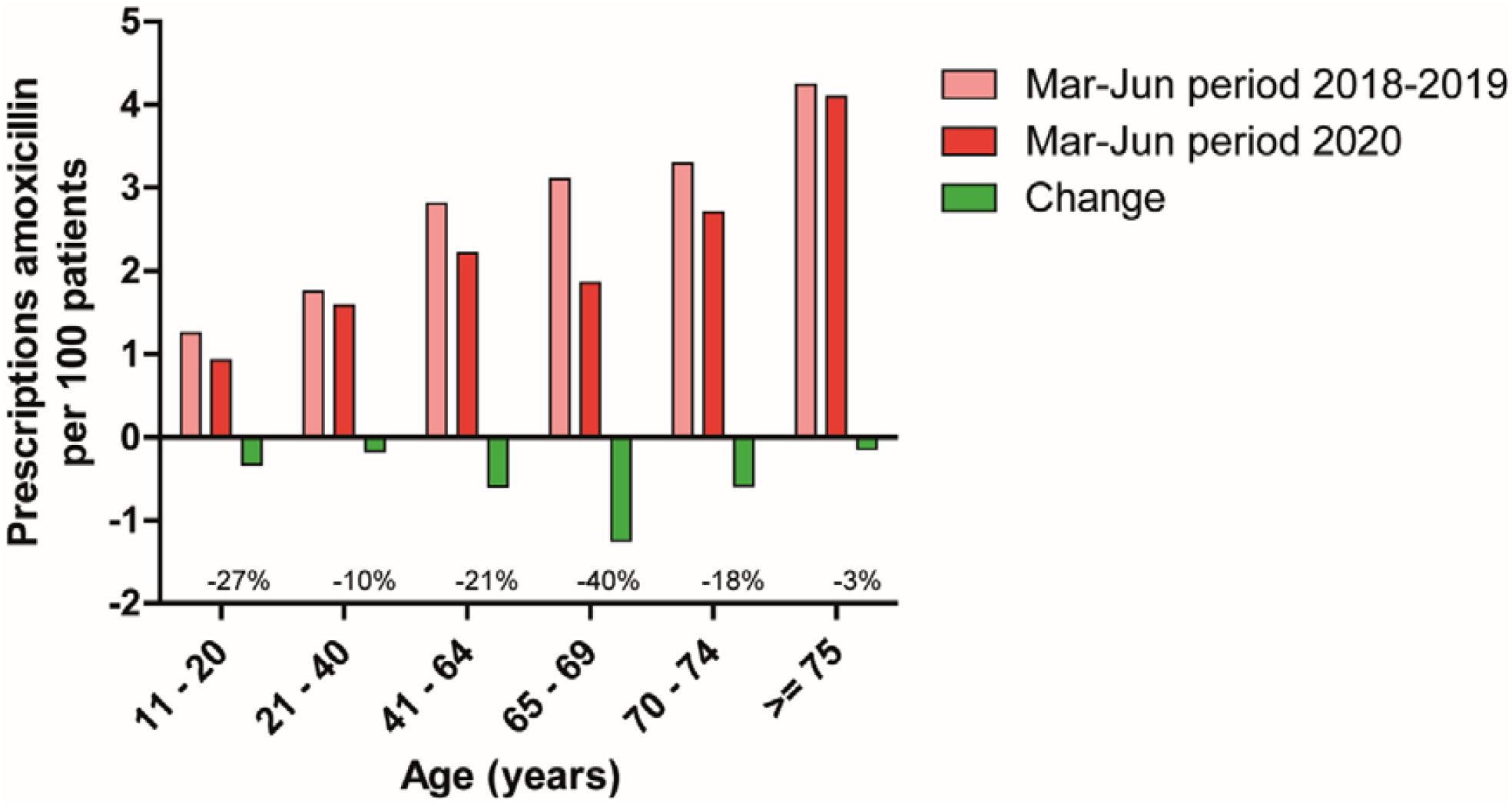
Decreases in prescription of amoxicillin by GPs in the study area during the first COVID-19 wave, stratified by patient age.

Infections by respiratory viruses like Influenza come with a serious risk of bacterial superinfection [19]. In our dataset however, only 1 out of 13 adult bacteremia IPD cases was simultaneously diagnosed with COVID-19. In line, while potential pathogens have been identified in nasopharyngeal specimens of COVID-19 cases [20] actual co-infections are infrequently observed [21, 22]. SARS-CoV-2 is a less potent inducer of proinflammatory cytokines compared to influenza [23]. Another explanation for the relatively low number of IPD and COVID-19 co-infections could be the fact that during the first COVID-19 wave adults mainly contracted SARS-CoV-2 infection from contacts other than children [24, 25]. So, outside the pediatric population, avoiding concurrent exposition to the main reservoir for bacterial respiratory pathogens like *S. pneumoniae*.

Taken together, while social distancing measures will surely have mitigated transmission of *S. pneumoniae*, a relevant amount of infections may still have occurred among adults.

### Restricted hospital referral

In the Netherlands, GPs are the primary consultant for medical issues and act as gatekeepers who determine if referral to the hospital or Emergency Department (ED) is indicated. While the number of GP consultations for respiratory infections peaked during our study period, in parallel patients frequently abstained from seeking medical care or abstained from in-hospital treatment out of fear of SARS-CoV-2 transmission, fear to overburden the health care system, and other personal grounds [26-28]. Once referral was desired and indicated GPs experienced no particular barriers towards ED evaluation during the first wave of COVID-19, yet only part of these evaluations led to hospitalization and microbiological diagnostic testing.

Outpatient management of bacteremic pneumococcal pneumonia with oral amoxicillin will often be reasonably adequate in the Netherlands, as pneumococci are generally susceptible. The risk of complications like pleural empyema is around 6% however and, in that case, (prolonged) antibiotic treatment without drainage may be suboptimal. Meningitis is a clinical emergency for which treatment with oral penicillins will most often fail. However, these specific complications can be hard to recognize in the outpatient setting. More generally, outcomes for elderly with moderate to severe disease are likely to be affected by restricted outpatient management. Despite these infection-oriented considerations, assuming that residual IPD has taken place, home treatment seems to have been a deliberate choice as hardly any delayed or complicated admissions with positive blood cultures have been observed.

Clinical diagnoses of the 13 adult pneumococcal bacteremia cases identified during the first COVID-19 wave in the Netherlands showed a regular distribution: 10 had a pneumonia (3 complicated with pleural effusion of which 1 proven empyema), and the other manifestations were 1 proven meningitis, 1 suspected meningitis without CSF culture performed, and 1 skin infection. However, several other characteristics suggest that a particular subset of IPD cases was hospitalized. In the months preceding the first COVID-19 wave obesity was markedly common among adult IPD patients (40%), while this proportion normalized from March onwards. Figure 4 and Table 1 demonstrate that identified IPD cases were referred without delay and generally concerned elderly women with comorbidity and severe pneumonia. The proportion of cases with cancer was twice as high compared to earlier years. Eight IPD cases with 5 different types of solid tumors and hematological malignancies were captured. Significantly fewer IPD patients were eligible for ICU-treatment during the first COVID-19 wave compared to the previous years (Table 2). Towards the end of the first COVID-19 wave, early cases in May concerned suspected meningitis cases and erysipelas irresponsive to antibiotic treatment. Only after the relief of certain social distancing measures, bacteremic pneumococcal pneumonia cases reemerged. Therefore, it seems that during the peak of COVID-19 only seriously ill elderly with notable comorbidity were selected for in-hospital treatment.

**Table 1:**
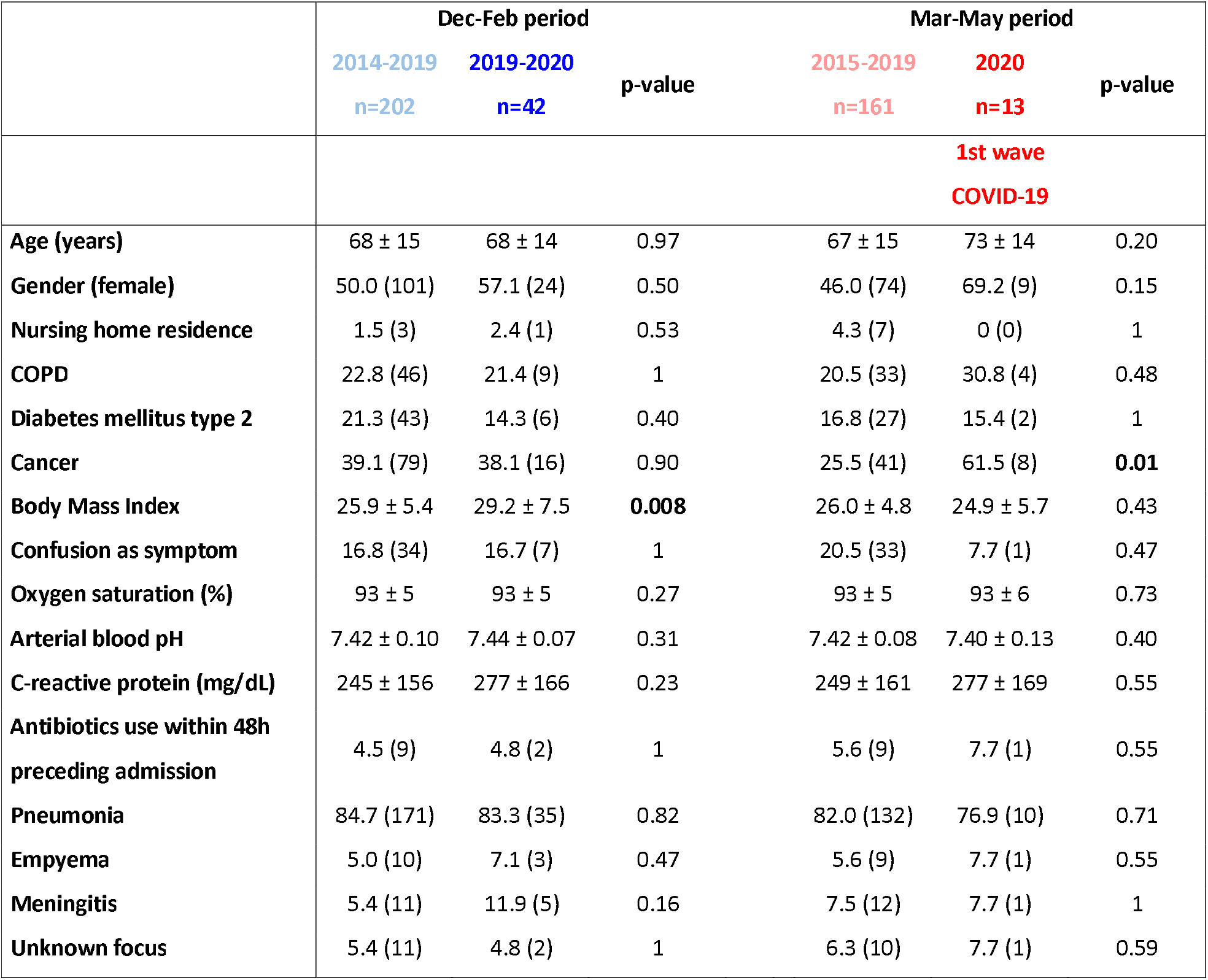
Clinical characteristics of adult IPD patients per time period

**Table 2:**
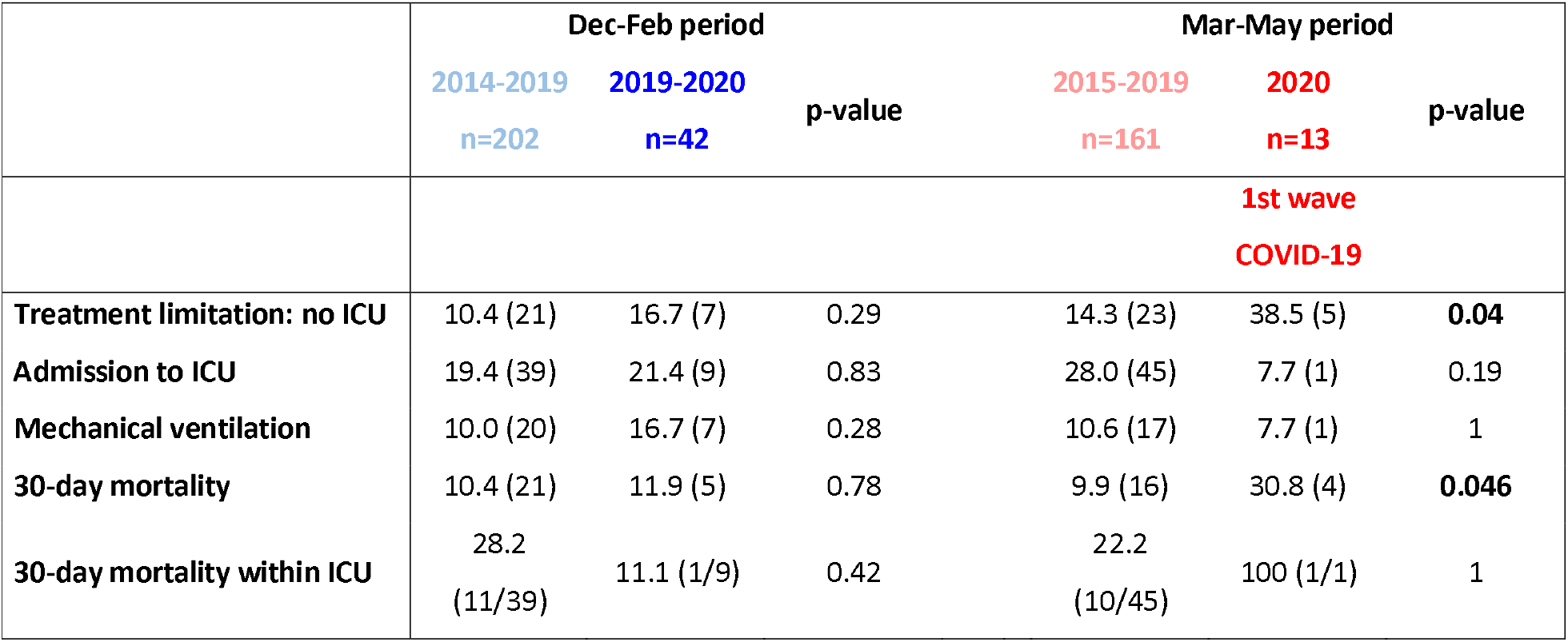
Aspects of clinical care for adult IPD patients per time period

**Figure 4:**
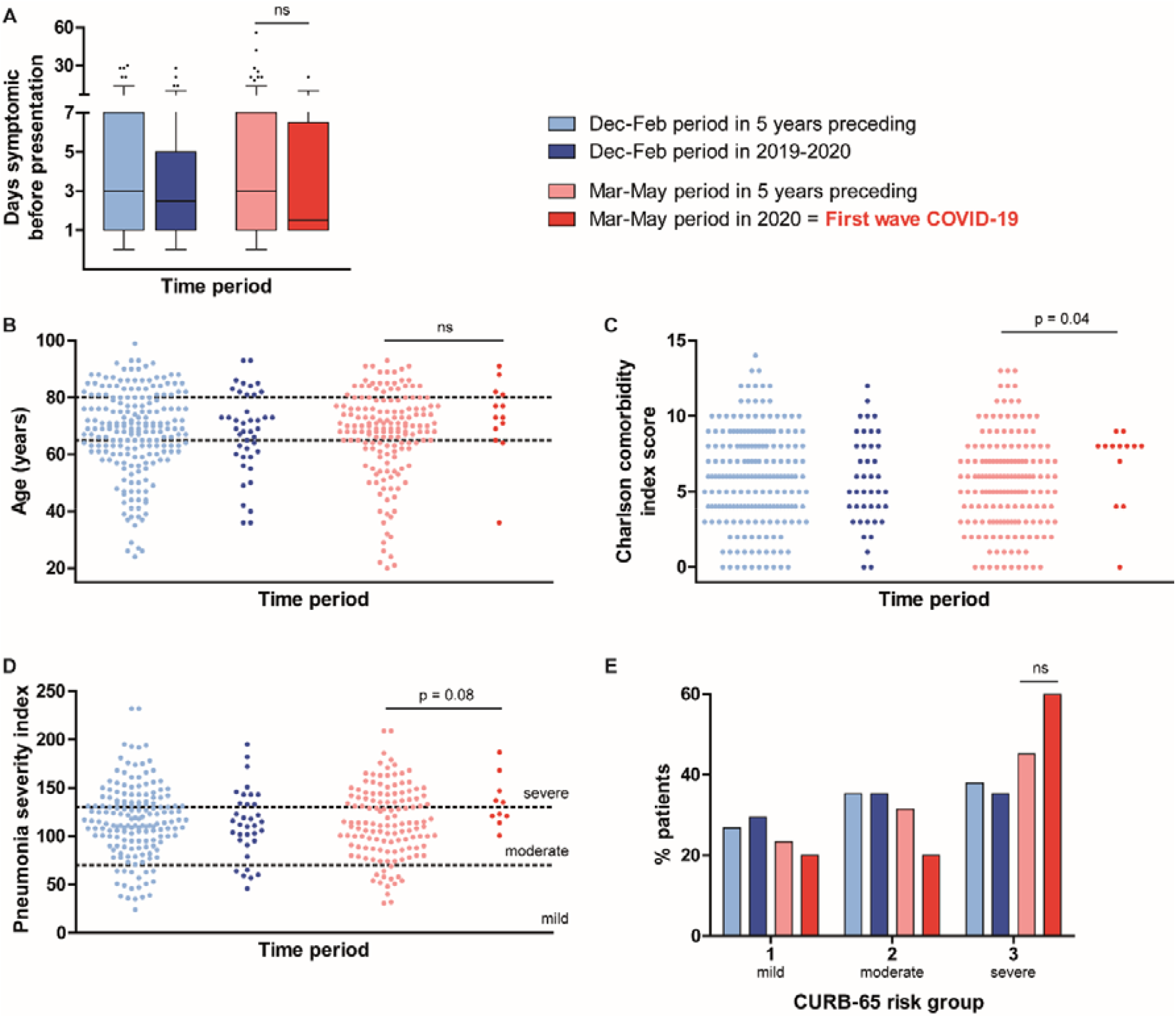
Patient characteristics of adult IPD cases capture during the first wave of COVID-19 (in red) compared to that time period in 5 years preceding (pink), and to the season preceding. Time to presentation at the hospital is displayed in panel A, baseline health is represented in panels B and C, and severity of pneumonia cases at the emergency department is shown in panels D and E.

Another interesting observation is the earlier decline in IPD cases in hospitals CWZ and Radboudumc compared to Rijnstate (Figure 1). The first 2 hospitals are located in Nijmegen ‘below the rivers’, referring to the dividing line between the North and South of the Netherlands formed by the rivers Waal and Rhine. The first wave of COVID-19 originated in the South of the Netherlands and made its way upwards consecutively occupying hospitals’ capacity. The observed order in reduction is an indication that restricted healthcare access has contributed to the drop in identified adult IPD cases.

### Impediments for diagnostic testing

During the first COVID-19 wave the benefit of hospital admission for patients with suspected respiratory infection was stringently assessed by both the GP’s and at the Emergency Department, and for patients who returned home generally no microbiological testing was performed. Although diagnostic capacity was under pressure, for hospitalized patients all 3 hospitals were able to maintain standard blood culture practices with respect to indication and incubation time. CWZ temporarily went from 4 to 2 blood culture bottles per patient as maximum incubator capacity was reached, yet for *S. pneumoniae* in 90% of cases both blood culture sets are positive [29]. At this hospital the number of adults from whom blood cultures were collected at the Emergency Department from March to May was 45% higher than the year before (915 in 2019 and 1,328 in 2020). This expansion in diagnostic effort was most prominent for patients 20-60 years old (92% higher), but also took place for patients over 75 (15% higher). Relative increases in blood cultures performed at the Emergency Department were more modest at Radboudumc (+ 8%) and Rijnstate (+ 18%). *S. pneumoniae* was cultured from 1.5% (14/915) versus 0.3% (4/1,328) of blood cultures performed at CWZ during the 2019 and 2020 periods respectively (p=0.018). Although rates of blood culture contamination were elevated during the first COVID-19 wave in all 3 hospitals, it is unlikely that this has masked the identification of *S. pneumoniae* as its time to blood culture positivity is generally short. Delays between collection blood cultures and start of incubation do affect chances of pneumococcal growth [30], but this has not been an issue in the participating hospitals with blood culture incubators present within the central hospital building.

Prior use of antibiotics heavily impedes pathogen identification in community acquired pneumonia (CAP), because it hampers bacterial growth in diagnostic cultures, also when the antibiotic may be unsuited to treat pneumococcal infections [10]. *S. pneumoniae* is especially sensitive to prior antibiotics use with odds for positive blood cultures dropping to 0.20, compared to 0.54 for all CAP-causing bacteria [31]. For the Dutch CAP population, antibiotics use within 14 days prior to hospital presentation is usually around 30% [32, 33]. However, a random sample of patients presenting with COVID-19 to the study hospitals showed that 40% had used antibiotics in the past 2 days, amoxicillin allocated to the majority. The high number of COVID-19 patients pre-treated with antibiotics is in contrast to the 13 identified adult IPD cases of whom only 1 had received antibiotics within 48 hours prior to presentation; long term doxycycline prophylaxis to which the pneumococcal isolate was resistant. Furthermore, cultures of cerebrospinal fluid are probably less affected by prior oral antibiotics use than blood cultures [34, 35]. Outside our study population of pneumococcal bacteremia, up to September 2020 already 3 cases of pneumococcal meningitis with negative blood cultures were established, while this normally occurs about once a year.

Therefore, we presume that more patients with pneumococcal bacteremia did reach the hospital, but have not been recognized as such due to prior use of antibiotics or because of return home without microbiological testing.

### Modifications in hospital care

To assess whether 30-day mortality among the 13 identified IPD cases actually deviated from expected, a case-control analysis was performed. Cases were adults with IPD in 2020, who were each matched to 4 controls from 2015-2019 based on baseline probability of dying from advent IPD (age, comorbidity, focus of pneumococcal infection). The relative risk of being infected during the first COVID-19 wave among deaths compared to survivors was 1.96 (95% confidence interval: 0.7 -5.3; p = 0.19). Therefore, mortality did not significantly deviate from expected in the IPD sample that was captured. At the same time, hospital care for IPD cases may have been modified during the first wave of COVID-19 due to isolation practices applied and increased public engagement in treatment restrictions.

Adult IPD cases were not generally notifiable during the study period, yet CWZ and Rijnstate both serve as sentinel laboratories for national IPD surveillance and reported all identified cases within 5 days after blood culture collection, also during the pandemic.

### Summarized

We studied changes in confirmed cases of adults with pneumococcal bacteremia during the first wave of COVID-19 in three adjacent hospitals in eastern Netherlands, compared to the five preceding years. We conclude that the drop in identified cases is multifactorial, with likely contributions of repressed referral practices and impaired diagnostic yield, in addition to decreased transmission (Table 3). Expected cases that were not captured compared to the years before are elderly men and the younger population with milder disease. Our data suggests that these patients may have gotten infected, but did no longer reach the hospital or were not identified due to prior use of antibiotics. The mortality among adult IPD cases that were identified was not higher than expected.

**Table 3:**
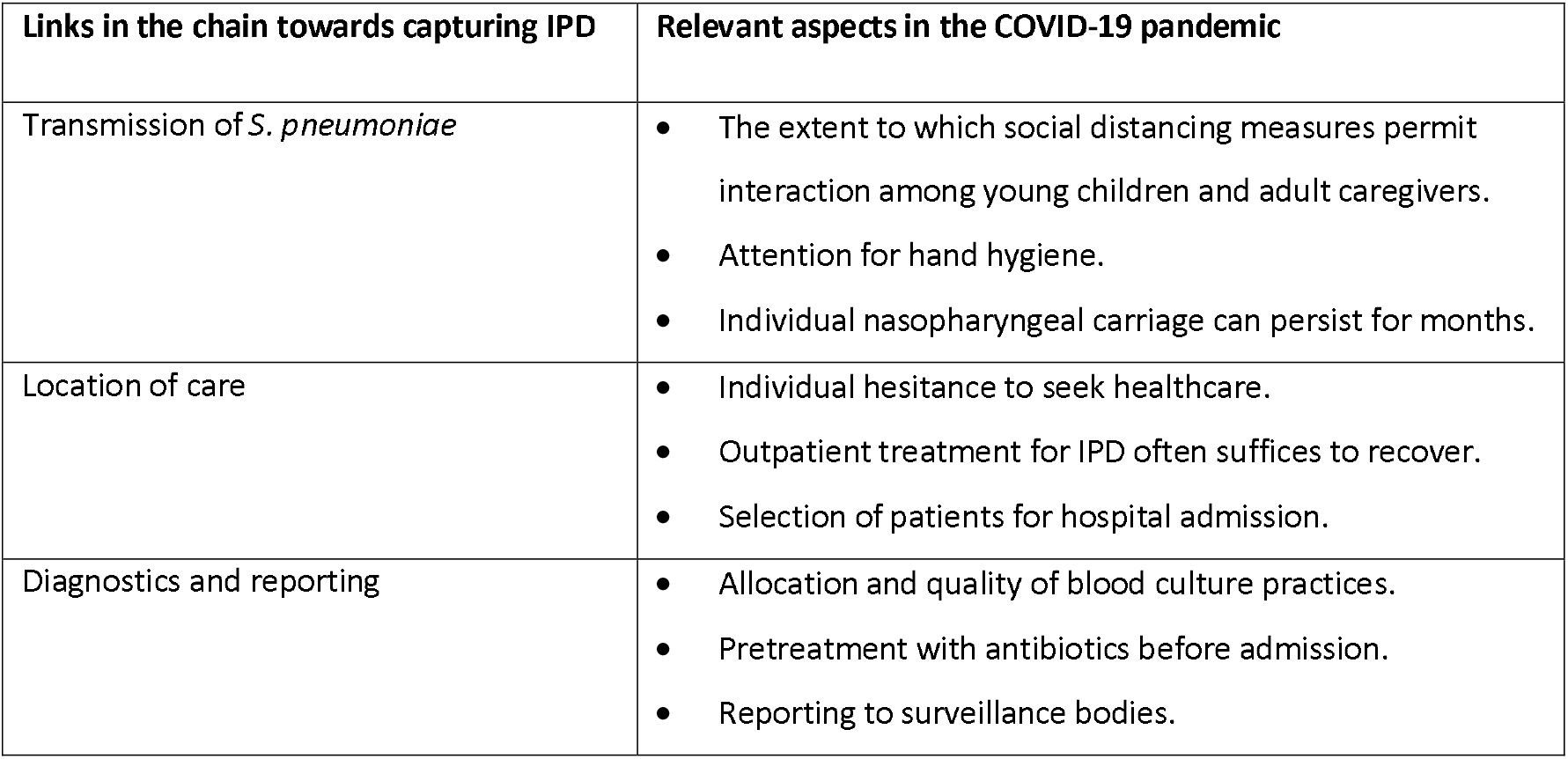
Summary of factors that contribute to the reporting of IPD.

| Links in the chain towards capturing IPD | Relevant aspects in the COVID-19 pandemic |
| --- | --- |
| Transmission of *S. pneumoniae* | • The extent to which social distancing measures permit interaction among young children and adult caregivers.<br>• Attention for hand hygiene.<br>• Individual nasopharyngeal carriage can persist for months. |
| Location of care | • Individual hesitance to seek healthcare.<br>• Outpatient treatment for IPD often suffices to recover.<br>• Selection of patients for hospital admission. |
| Diagnostics and reporting | • Allocation and quality of blood culture practices.<br>• Pretreatment with antibiotics before admission.<br>• Reporting to surveillance bodies. |

### Evaluation and possible solutions

Illustrative for transmission being just one of multiple links in the chain towards capturing IPD, is that 7 out of 8 bacteremic IPD cases with solitary pneumonia in our study had cancer. These patients are more readily admitted to the hospital during the pandemic directly by their attending medical oncologist, without interference of a GP or prior antibiotics, which increases the chances of IPD being detected. Patients with presumed pneumonia, their GPs, and their assessors at the ED have been more critical towards the desire for in-hospital treatment, and often opted for treatment at home in void of microbiological testing. Such selection bias will flaw surveillance on the level of incidence estimates, but it may also affect the validity of antimicrobial resistance and serotype dynamics.

International IPD surveillance data demonstrated that, after a sharp drop in mobility, IPD incidence rates kept gradually decreasing over months [36]. This additional attenuation of expected cases could not be attributed to school closures. Also, the transmission of respiratory viruses like influenza that may predispose for IPD ceased instantaneously after the Dutch lockdown, not tapering off like IPD [37, 38]. It may rather represent universal gradual waning of pneumococcal colonization and subsequent disease. Instant drops in IPD reporting are more likely to be related to changes health care provision and diagnostic yield, and therefore more country-specific. In the Netherlands where pneumococci are susceptible to first line antibiotics and a strong GP-system is in place, the instant decline in captured IPD was probably not just attributable to decreased transmission.

Of particular interest is the comparison between *S. pneumoniae* and *Haemophilus influenzae*. These two respiratory pathogens have similar transmission and disease dynamics in the population, yet the incidence of captured invasive *H. influenzae* infections among adults has not decreased during the pandemic in the Netherlands [39]. The discordance to IPD may again be explained by particular Dutch treatment and referral practices. *H. influenzae* is more steadily cultured [40], more often resistant to amoxicillin, and the population at risk for invasive *H. influenzae* infection are adults with humoral immunodeficiencies who are often already in specialist care with hospitalization and microbiological diagnostic testing more readily performed.

To maintain sight on IPD dynamics there may be ways that are less affected by (future) fluctuations in clinical practices. It could include sampling at alternative health care providers, or using alternative diagnostic techniques. Global surveillance of pneumococcal serotypes in IPD is currently based on blood and cerebrospinal fluid cultures. Although the use of pneumococcal urinary antigen tests is under debate because of limited sensitivity (60-75%) and protracted positivity, their results are not affected by prior antibiotics use and could be generated outside the hospital [41]. Moreover, sensitivity issues can be overcome using a CRP threshold above which urinary antigen testing will be representative [42, 43]. Also, detection of pneumococcal DNA in blood specimens could be an alternative to establish the presence of pneumococcal bacteremia as the DNA remains detectable for days after initiation of antibiotics and is absent in healthy carriers of *S. pneumoniae* [44, 45]. This diagnostic modality yields additional microbiological diagnoses in CAP patients, especially in cases with prior use of antibiotics in which other tests failed to identify *S. pneumoniae* [46]. For treatment decisions in the GP setting, it is unsure whether the result of either test (urinary antigen and blood DNA) would make a difference. The particular benefit for surveillance purposes is that both can be performed in retrospect on stored specimens. In addition, genotyping pneumococcal DNA from blood can provide information on serotype, virulence factors, and antibiotic resistance markers [47, 48]. In the Netherlands shifts in serotype distribution are anticipated given the nationwide roll out of polysaccharide pneumococcal vaccination in the elderly population. As culture is the gold standard for invasive pneumococcal infections it is an option to retain hospital-generated culture data as primary source for monitoring vaccine impact. However, in areas where referral and diagnostic practices are unstable, other diagnostic techniques may provide a valuable alternative to monitor incidence and serotype dynamics of IPD, even in patients who are not hospitalized.

Inherent to the analysis of a reduced number of cases is that the evidence presented in this study is based on inference. Our theory of undetected residual IPD cases desires confirmation by alternative study design. Furthermore, this study was limited to a confined region and to adults with positive blood cultures. Therefore, children and rarer cases of IPD with *S. pneumoniae* cultured solely from a different site of infection were not included. For adults however, we think that our in-depth analysis beyond the reported numbers provides a relevant impression and a useful framework to assess potential causes of decreased reporting of infectious diseases like IPD in times of a pandemic.

## Data Availability

Data available upon request and in accordance with approved study procedures.

## Acknowledgements

This study was initiated by the BACON study group (Bacteremia Collection East Netherlands) as part of a research project on Genomic epistasis of invasive *S. pneumoniae* funded by a Research Grant 2020 from the European Society of Clinical Microbiology and Infectious Diseases (ESCMID) to A.J.H. Cremers. We acknowledge all participating hospitals and affiliated researchers for their contribution and support. We thank pharmacist Sjanne van Roijen for sharing the data on antibiotic prescriptions in the GP population.

## Methods

### Study population and data collection

IPD data were retrospectively obtained from a multicentered cohort study using clinical and microbiological digital patient systems from three hospitals in the East of the Netherlands: Canisius-Wilhelmina Ziekenhuis (CWZ) Nijmegen, Radboudumc Nijmegen and Rijnstate Ziekenhuis Arnhem. All hospitalized patients aged 18 and older with microbiologically proven *S. pneumoniae* bacteremia admitted between January 1^th^ 2012 until July 1^th^ 2020 were considered.

The collected data included basic demographic data, comorbidities, clinical data including signs and symptoms, duration of disease, prior use of antibiotics and outcome, and diagnostic information such as laboratory analysis, microbiological analysis, and radiography. Limitations to treatment, either patient initiated of medically advised (mechanical ventilation and ICU treatment) was also recorded for each patient.

### Microbiological analysis

At the emergency department two sets of blood cultures were sampled. Patients developing fever during admission had one set of blood cultures sampled. Blood cultures were processed according to standard microbiological procedures. Isolates were identified using malditof. Serotypes were provided by the Dutch Reference Laboratory Bacterial Meningitis (NRLBM).

### Data pertaining the factors considered

Government legislation affecting exposure to possible carriers was gathered from government websites and we compared the ensuing timeline to the observed decrease in IPD cases. Changes in prescribed antibiotics were calculated using data over years 2018, 2019 and 2020 from local pharmacies. To assess changes in blood culture processing capacity a database entailing all blood cultures sampled at one of the hospitals between January 2019 and July 2020 considering a single blood culture per person per febrile episode was analyzed. Consecutive blood cultures sampled at least 14 days after initial blood cultures were considered separate episodes and therefore included in analysis.

### Data management and analysis

Clinical information was converted into an anonymous digital database, Castor EDC, for analytical purposes. The updated Charlson Comorbidity index (uCCI) was calculated for each patient.

Statistical analysis was performed using SPSS (IBM SPSS Statistics, Version 26.0). Patients were divided into groups based on time of presentation. The COVID-19 epidemic was defined as the time period between March 1^th^ and May 31^th^ 2020. As appropriate, differences in time periods were assessed using Chi-square test, Fisher-Exact test, one-way-ANOVA and Kruskal-Wallis test. A case-control study was performed to assess changes in 30-day mortality. All statistical tests were performed two-sided and a p-value of < 0.05 was considered statistically significant.

### Ethical considerations

The study was approved by a central Medical Ethics Committee as well as the local Ethics Committees of all participating hospitals.

